# APOE *ε*4-Associated Hippocampal Atrophy Trajectories Across the Alzheimer’s Disease Continuum: A Systematic Review, Meta-Analysis, and Longitudinal Validation

**DOI:** 10.64898/2025.12.08.25341534

**Authors:** Minnuo Cai, Hang Lei, Yuetong Zhang, Jiaxiang Zou, Wanjing Cao, Yiquan Wang, Kai Wei, the Alzheimer’s Disease Neuroimaging Initiative

## Abstract

**Background:** The role of APOE-*ε*4 in hippocampal atrophy is contested. We aimed to determine whether it functions as a static risk factor or an amyloid-*β* (A*β*)-dependent modulator of neurodegeneration.

**Methods:** We integrated a systematic meta-analysis of 18 studies (*N* = 3, 781) with longitudinal validation using linear mixed-effects models in the NACC and ADNI cohorts (*N >* 5, 000), employing biomarker stratification to test for gene-pathology interactions.

**Results:** The meta-analysis confirmed significant atrophy in APOE-*ε*4 carriers but with high heterogeneity. Longitudinal analysis resolved this by identifying a crucial interaction: in A*β*-negative individuals, carrier atrophy rates were indistinguishable from non-carriers. However, A*β* positivity triggered a dramatic, dose-dependent acceleration in atrophy among carriers, with homozygotes declining over three times faster.

**Conclusions:** APOE-*ε*4 acts as a potent, conditional accelerator of neurodegeneration, not an independent driver. Its deleterious effect is contingent on the presence of A*β* pathology. Clinical risk stratification should therefore integrate amyloid status with APOE genotype to accurately predict structural progression.

## 1 Introduction

Alzheimer’s Disease (AD) is biologically defined by the accumulation of amyloid-*β* (A*β*) and tau proteins, and characterized by progressive hippocampal atrophy as a key marker of neurodegeneration [1–3]. While the *ε*4 allele of Apolipoprotein E (APOE-*ε*4) is recognized as the strongest genetic risk factor for late-onset AD [4,5], the precise nature of its impact on hippocampal integrity remains incompletely understood.

Specifically, critical knowledge gaps persist regarding the timing, mechanism, and dosage of APOE-*ε*4-related atrophy. First, conflicting evidence exists regarding the temporal origin of structural loss: the developmental hypothesis suggests a static, congenital phenotype [6, 7], whereas the neurodegenerative hypothesis proposes an accelerated decline in later life [8–10]. Second, the allele dosage effect, specifically whether homozygous (*ε*4/*ε*4) carriers exhibit disproportionately severe atrophy compared to heterozygotes, is not fully characterized, leaving the linearity of potential genetic toxicity uncertain [11, 12]. Third, a qualitative divergence exists regarding whether APOE acts through direct structural toxicity [13] or primarily as an upstream modulator dependent on amyloidosis [9].

These unresolved discrepancies likely stem from both biological and methodological limitations in prior literature. Biologically, traditional volumetric analyses often overlook occult pathologies, such as asymptomatic amyloidosis, in clinically normal populations [14, 15]. This may confound independent genetic effects with early-stage disease processes. Methodologically, previous studies have been constrained by modest sample sizes, varying measurement protocols, and demographic heterogeneity [16]. This variability limits statistical power and hinders a robust, consensus-based assessment of the APOE effect size across the AD continuum.

To address these gaps, this study employs a comprehensive, hierarchical design moving from macro-level synthesis to mechanistic analysis. We first conduct a systematic meta-analysis to integrate large-scale data, overcoming individual study heterogeneity to quantify the aggregate effect of APOE-*ε*4. Subsequently, we utilize dual-cohort longitudinal validation (NACC and ADNI) with biomarker stratification [17,18]. This integrated approach aims to elucidate whether APOE-*ε*4-related atrophy represents a dose-dependent, intrinsic feature or a conditional phenomenon associated with the synergistic interaction of amyloid pathology [9].

## 2 Methods

### 2.1 Meta-analysis Strategy

This systematic review and meta-analysis was conducted and reported in accordance with the PRISMA guidelines [19] (Figure 1). We performed a systematic search of PubMed, Embase, the Web of Science Core Collection, and the Cochrane Library for original research published up to August 2025, following the Population, Intervention, Comparison, and Outcome (PICO) framework. The entire screening process was managed using the Covidence online platform.

**Figure 1.**
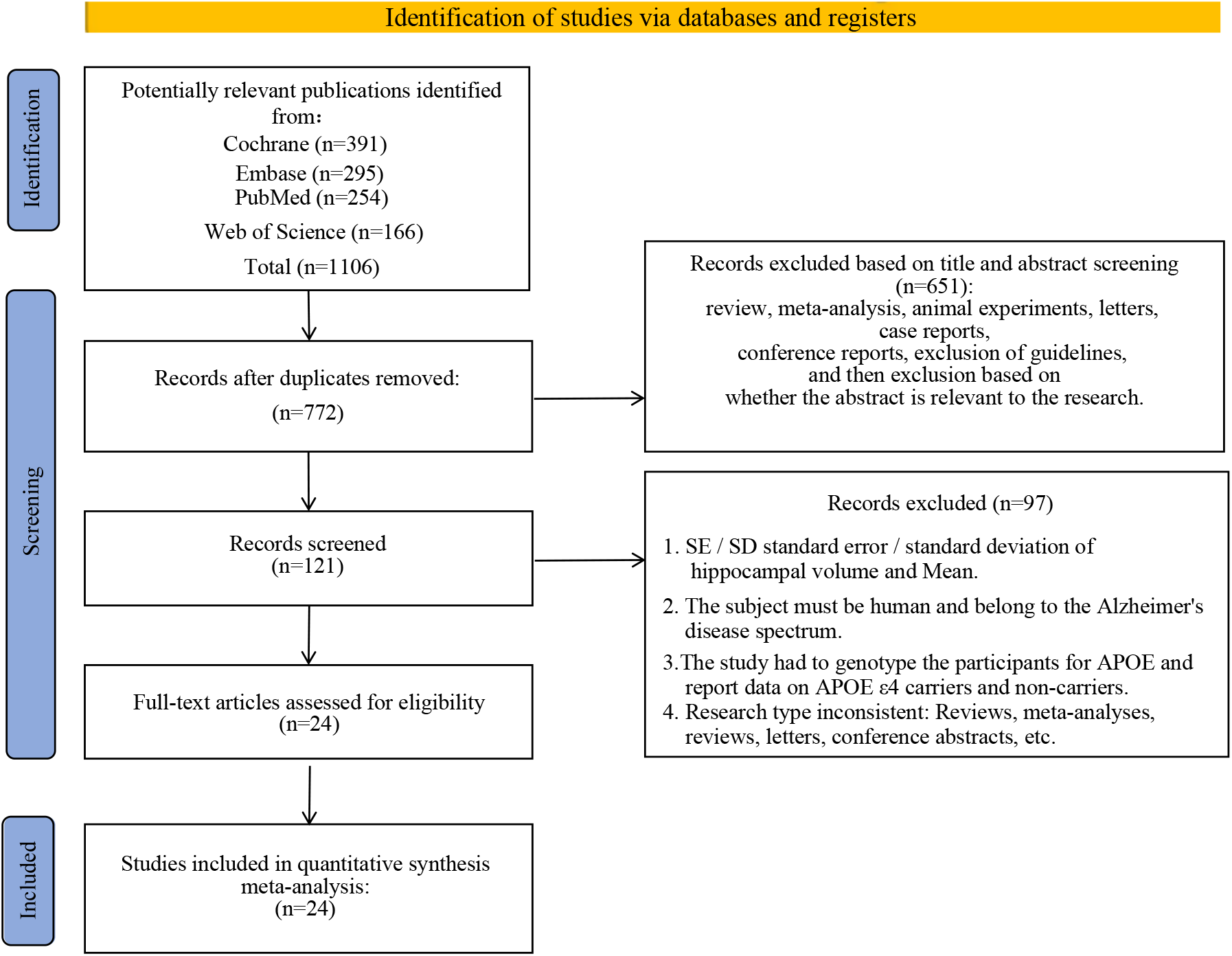
PRISMA flow diagram of the study selection process. The flowchart depicts the stepwise exclusion of records from identification to final inclusion. A total of 1,106 records were initially identified from four databases (Cochrane, Embase, PubMed, and Web of Science). Following duplicate removal, 772 records underwent title and abstract screening. Subsequently, 121 full-text articles were assessed for eligibility, with 97 excluded for not meeting the predefined inclusion criteria (see Supporting Information for details). Ultimately, 24 studies met all inclusion criteria for the quantitative meta-analysis.

A notable challenge in this field is the use of overlapping data from large, shared databases across multiple publications. To address this, we implemented a specific protocol: if multiple studies originated from the same primary data source, we included only the publication that was most recent or provided the largest and most comprehensive sample. This step was essential to ensure the independence of the included cohorts and refined our initial selection from 24 eligible articles (*N* = 7326) to a final set of 18 unique studies (*N* = 3781) [6, 20–42]. For the quantitative synthesis, we extracted sample sizes, means, and standard deviations of hippocampal volume for both APOE-*ε*4 carriers and non-carriers from these publications.

The effect size was calculated as the Standardized Mean Difference (SMD) using Hedges’ g. A random-effects model, with the between-study variance estimated using Restricted Maximum-Likelihood (REML), was employed to pool effect sizes [43]. The Hartung-Knapp adjustment was applied for more robust confidence intervals. Heterogeneity was quantified using the *I*^2^ statistic [44]. To investigate sources of heterogeneity, we first performed subgroup analyses based on clinical diagnosis, APOE genotype dosage, and ICV correction methods [45]. Second, we conducted univariable random-effects meta-regressions to assess the potential moderating effects of mean participant age and sex distribution (Figure 2).

**Figure 2.**
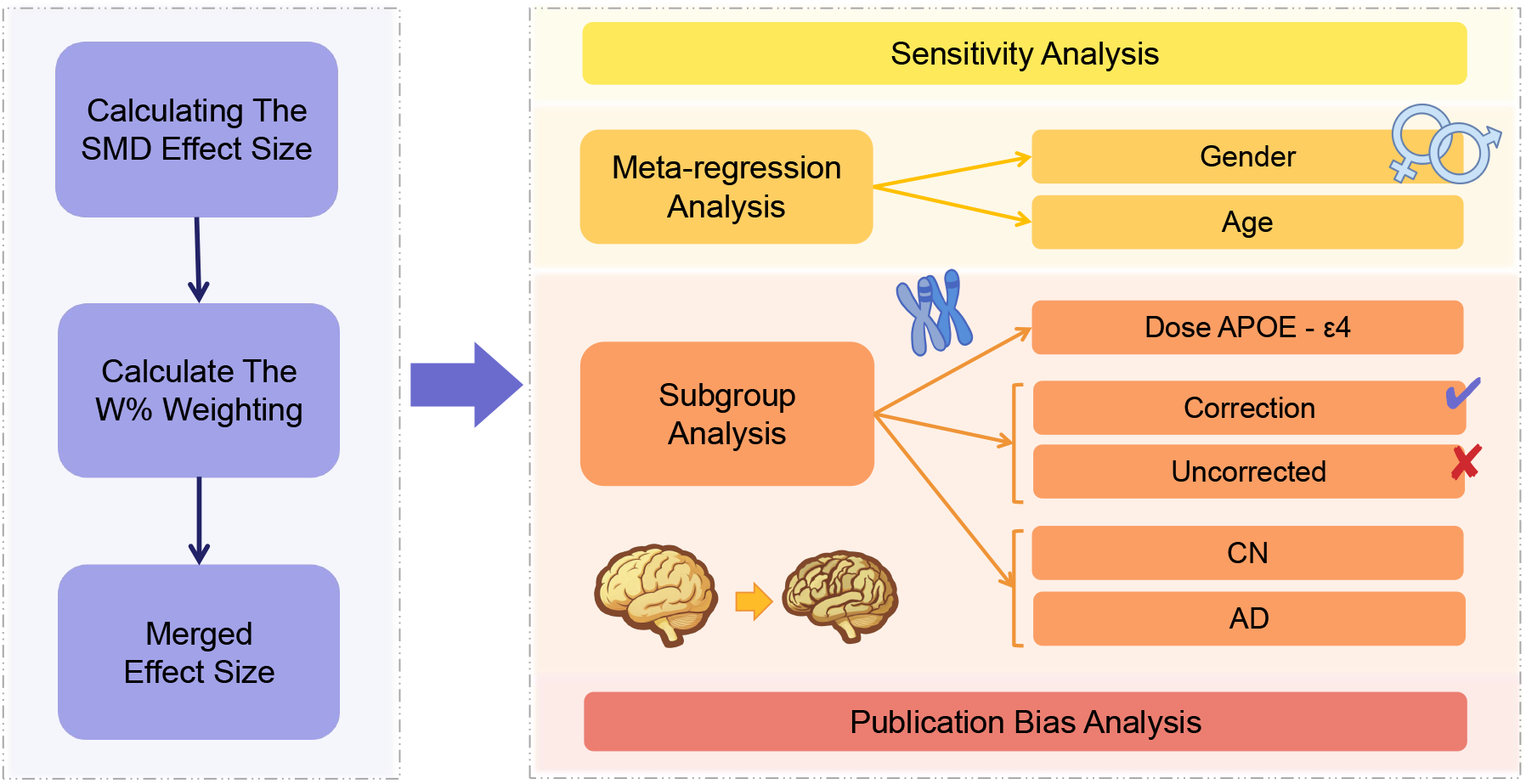
Schematic framework of the quantitative synthesis and heterogeneity exploration. The left panel depicts the sequential procedure for pooling effect sizes, moving from individual Standardized Mean Difference (SMD) calculation to weight assignment (W%) and the final merged estimation. The right panel details the strategies employed to investigate heterogeneity, including sensitivity analysis and meta-regression (assessing Gender and Age). Subgroup analyses are stratified by three key domains: APOE-*ε*4 genotype dosage, methodological correction (Corrected vs. Uncorrected), and clinical stage (Cognitively Normal, CN; AD). Publication bias assessment is performed as the final validation step.

To evaluate the stability and robustness of our results, several additional analyses were conducted. We performed a leave-one-out sensitivity analysis to assess the influence of each individual study on the overall pooled estimate. Baujat plots were generated to visually identify studies contributing most to both overall heterogeneity and the pooled effect. Furthermore, we implemented a multiverse analysis to test whether the main conclusion was robust to different analytical choices regarding the removal of potential outliers. Finally, publication bias was assessed using Egger’s regression test, and the trim-and-fill method was used to estimate a bias-corrected effect size [46]. All statistical analyses were performed in R (version 4.5.2).

### 2.2 Longitudinal Cohorts and Participants

We utilized longitudinal data from two independent multicenter cohorts: the National Alzheimer’s Coordinating Center (NACC) and the Alzheimer’s Disease Neuroimaging Initiative (ADNI).

The NACC Uniform Data Set was utilized to assess the gene-dose effect in a large-scale clinical population [18, 47]. Using the dataset from the September 2025 data freeze (covering visits between September 2005 and September 2025), we identified 3,986 participants who had available pre-processed longitudinal hippocampal volume measurements and confirmed APOE genotyping.

The ADNI dataset was employed to investigate the interaction between genetics and pathology [48, 49]. From the data downloaded in August 2025, we selected a subset of 1,947 participants based on the concurrent availability of longitudinal hippocampal volume measurements, confirmed APOE genotyping, and baseline Cerebrospinal Fluid (CSF) biomarker levels (A*β*42, p-Tau181, and t-Tau). No raw MRI images were processed in this study; all volumetric data were obtained directly from the study repositories.

To ensure the independence of the discovery and validation cohorts, systematic cross-verification was performed to confirm that there were no overlapping subjects between the two datasets based on unique identifiers and demographic profiles.

### 2.3 Data Preprocessing and Quality Control

For the NACC dataset, a dedicated preprocessing and quality control (QC) pipeline was applied. First, to prevent endogeneity issues arising from disease progression itself, each subject’s clinical diagnosis was locked to their baseline status. Second, a two-stage QC was performed: for cross-sectional QC, we fitted a linear model of hippocampal volume adjusted for age, sex, eTIV, and baseline diagnosis, and observations with standardized residuals exceeding a conservative threshold of ±4 SD were excluded. For longitudinal QC, we calculated the annualized rate of volume change and removed trajectories with biologically implausible changes, such as an annual volume increase greater than 20% or a loss exceeding 30%. Finally, to optimize model performance and interpretability, the age covariate was centered and eTIV was scaled.

To construct a robust dataset for longitudinal analysis, a systematic preprocessing and quality control (QC) pipeline was applied to the ADNI data. To mitigate potential confounding from disease progression, each subject’s clinical diagnosis was fixed to their baseline status. The analysis was restricted to participants with at least two valid neuroimaging follow-up visits, ensuring the reliability of individual atrophy rate estimation. Subsequently, we identified and removed potential outliers by calculating the standardized Z-score for hippocampal volume and excluding observations where the absolute Z-score exceeded 4, a step intended to reduce potential interference from factors such as image segmentation errors. Following this filtering process, hippocampal volumes were re-standardized based on the final analytical sample for use in subsequent statistical models.

Additionally, to ensure the independence of the discovery and validation cohorts, we performed a systematic cross-verification between the ADNI and NACC ADC datasets. We confirmed that there were no overlapping subjects between the two datasets utilized in this study based on unique identifiers and demographic profiles.

### 2.4 Neuroimaging and Biomarker Definitions

This study utilized pre-computed hippocampal volume data provided by the NACC and ADNI databases. According to the protocols of these repositories, volumetric measures were derived from high-resolution T1-weighted MRI scans using automated segmentation pipelines (primarily FreeSurfer) [17, 50, 51].

For the ADNI longitudinal dataset, images were acquired using both 1.5T and 3T scanners and processed using FreeSurfer versions v4.4, v5.1, or v6.0 depending on the study phase. Specifically for the data utilized in this analysis, hippocampal volumes were adjusted for intracranial volume (ICV) to correct for inter-individual head size differences.

In contrast, the NACC dataset exclusively utilized scans ac-quired at 3T, processed with the more recent FreeSurfer v7.2. NACC volumes were adjusted using the estimated total intracranial volume (eTIV).

For the ADNI cohort, biomarker positivity was classified using the specific cut-off values provided in the dataset: A*β*42 *<* 976.6 pg/mL (Amyloid+), p-Tau181 *>* 21.8 pg/mL (p-Tau+), and t-Tau *>* 245 pg/mL (t-Tau+).

### 2.5 Statistical Models

Longitudinal changes in hippocampal volume were analyzed using Linear Mixed-effects Models (LMMs) [52, 53]. To account for within-subject correlations and individual heterogeneity in baseline volume and atrophy rates, all models included random intercepts and random slopes for time at the subject level.

#### Model 1: Gene-Dose Effect and its Modulators (NACC)

To quantify the dose-dependent acceleration of atrophy associated with APOE-*ε*4 and to explore potential modulation by age or sex, we modeled the hippocampal volume trajectories as follows:

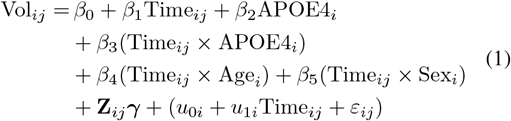

Here, Vol_*ij*_ represents the hippocampal volume for subject *i* at time *j*. The coefficient *β*_3_ captures the additional annual atrophy rate specific to *ε*4 carriers relative to non-carriers. The added interaction terms, *β*_4_ and *β*_5_, test whether atrophy rates vary with age or differ by sex, respectively. The term **Z**_*ij*_***γ*** denotes the main effects of covariates, including Age, Sex, Intracranial Volume (ICV), and Clinical Diagnosis.

#### Model 2: Specificity of Synergistic Interaction (ADNI, Individual Models)

To separately test for a synergistic interaction between APOE-*ε*4 and each core pathology in the ADNI cohort, we constructed a series of hierarchical models. The general form for a given biomarker (Bio_*k*_) is:

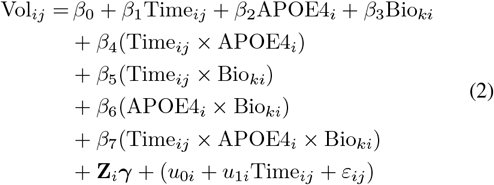

Here, APOE4_*i*_ is a binary indicator for carrier status, and the three-way interaction coefficient *β*_7_ is the primary term of interest.**Z** represent all main effects, two-way interactions, and covariates.

#### Model 3: Hierarchy of Synergistic Interaction (ADNI, Joint Model)

To assess the relative influence of A*β* and p-Tau pathologies, we constructed a joint model that simultaneously included all lower-order terms and the three-way interactions for both pathways:

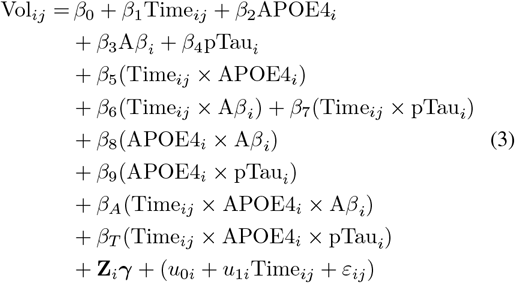

This model allows for a direct comparison of the interaction coefficients *β*_*A*_ and *β*_*T*_ after mutual adjustment.

#### Mediation Analysis (ADNI)

Given that initial model results suggested the effect of APOE-*ε*4 might be primarily associated with the A*β* pathway, a mediation analysis was conducted on the ADNI APOE-*ε*4 carriers. This analysis aimed to clarify the mechanistic relationship between A*β* and p-Tau. First, individual annual atrophy rates were calculated for each carrier. Subsequently, a mediation model was constructed with A*β* positivity as the independent variable (*A*), baseline p-Tau level as the mediator (*M*), and atrophy rate as the outcome (*Y*), while controlling for covariates. The significance of the indirect effect was assessed using the Sobel test.

## 3 Results

### 3.1 Meta-analysis: Overall Effect and Sources of Heterogeneity

Our quantitative synthesis included 18 eligible studies, which provided 26 distinct cohorts and a total of 4,311 participants. The random-effects meta-analysis showed a significant overall reduction in hippocampal volume for APOE-*ε*4 carriers compared to non-carriers (Standardized Mean Difference [SMD] = -0.27, 95% CI [-0.43, -0.11], *p* = 0.0017). The analysis indicated substantial heterogeneity across studies (*I*^2^ = 61.9%, *p <* 0.0001), suggesting that the pooled estimate reflects an average effect influenced by underlying moderators and warranting detailed subgroup analyses (Figure 3)(Table 1).

**Table 1.** Summary of Main Meta-Analysis Results.

| Analysis | k | SMD | 95% CI | P-value | $I^2$ | $\tau^2$ |
| --- | --- | --- | --- | --- | --- | --- |
| Overall | 26 | -0.27 | [-0.43, -0.11] | 0.002 | 61.9% | 0.077 |
| Subgroup: AD | 7 | -0.62 | [-0.94, -0.29] | 0.004 | 34.4% | 0.058 |
| Subgroup: CN | 18 | -0.16 | [-0.34, 0.02] | 0.071 | 55.3% | 0.054 |
| Method: Corrected | 14 | -0.41 | [-0.67, -0.16] | 0.004 | 74.0% | 0.123 |
| Method: Uncorrected | 12 | -0.10 | [-0.25, 0.06] | 0.186 | 24.1% | 0.002 |
| Bias Corrected (Trim-and-Fill) | 34 | -0.10 | [-0.30, 0.10] | 0.308 | 71.4% | 0.190 |

Note: k = number of studies; SMD = Standardized Mean Difference; 95% CI = 95% Confidence Interval; $I^2$ = heterogeneity; $\tau^2$ = between-study variance.

**Figure 3.**
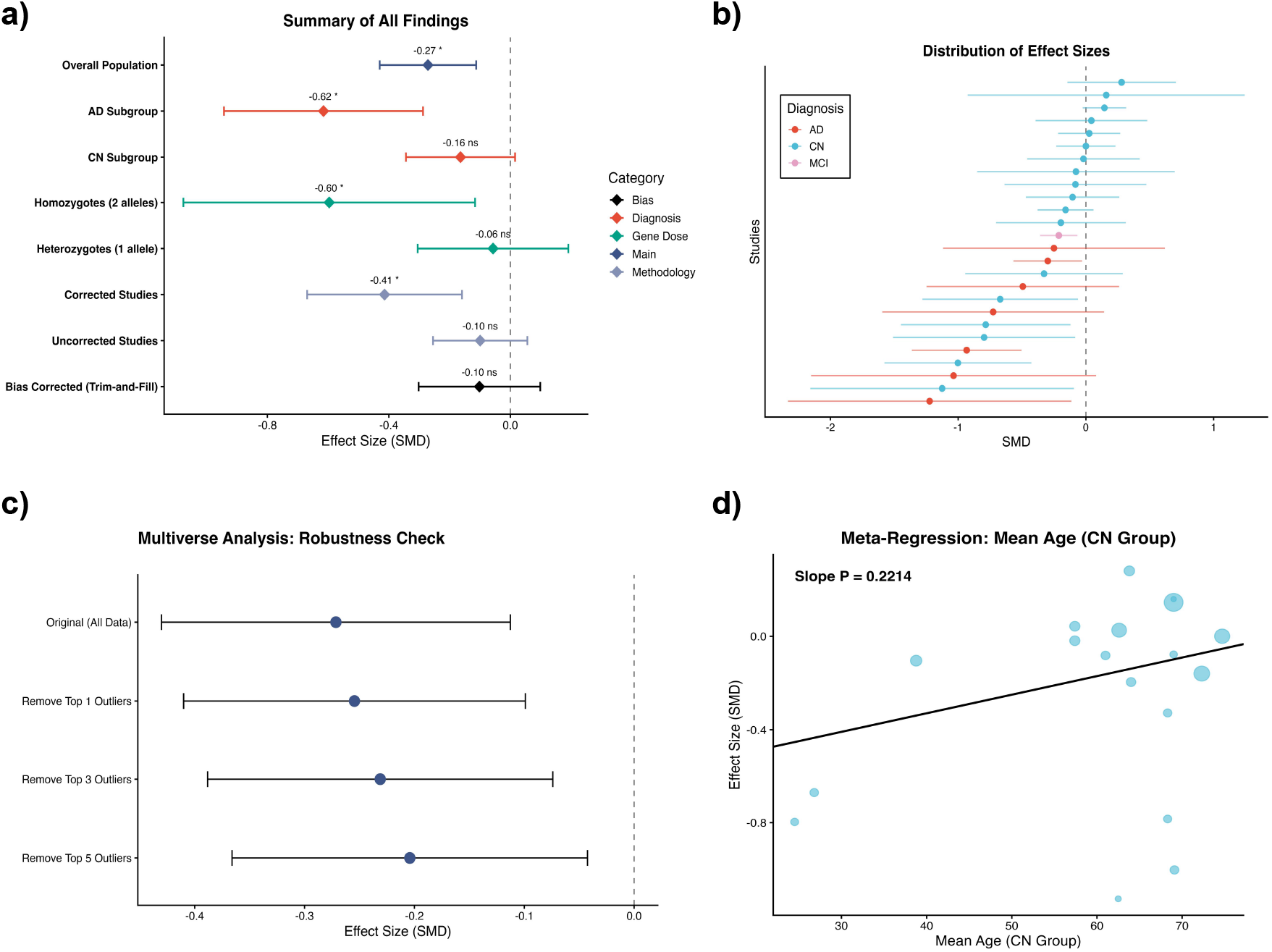
Meta-analysis of the association between APOE-*ε*4 and hippocampal volume. (a) Summary forest plot showing the overall pooled Standardized Mean Difference (SMD) and results from key subgroup analyses based on clinical diagnosis (AD, CN), genotype dosage, and methodological correction. A bias-corrected estimate using the trim-and-fill method is also presented. (b) Forest plot illustrating the distribution of effect sizes from all individual studies included in the analysis, color-coded by diagnosis. (c) Multiverse analysis as a robustness check, demonstrating that the overall conclusion remains stable after the sequential removal of the most extreme studies. (d) Meta-regression plot for the Cognitively Normal (CN) subgroup, assessing the moderating effect of mean participant age on the SMD. The non-significant slope (p=0.2214) suggests that age does not significantly explain the heterogeneity in this subgroup. In all forest plots, points/diamonds represent SMDs and horizontal lines denote 95% Confidence Intervals (CIs).

Stratification by clinical diagnosis indicated a pronounced gradient in the magnitude of APOE-*ε*4-associated atrophy, suggesting that disease stage is a key modulator of the effect size. The association was most pronounced in patients with Alzheimer’s Disease, who showed a large and significant volume reduction (SMD = -0.62, 95% CI [-0.94, -0.29], *p* = 0.0037), with reduced heterogeneity in this subgroup (*I*^2^ = 34.4%). In contrast, the effect in Cognitively Normal (CN) individuals was small and not statistically significant (SMD = -0.16, 95% CI [-0.34, 0.02], *p* = 0.071), with considerable remaining heterogeneity (*I*^2^ = 55.3%). This high variability suggests the influence of unmeasured factors, such as the varying prevalence of preclinical amyloid pathology across CN cohorts. We hypothesized that older age, as a proxy for higher amyloid risk, might moderate this effect. However, a specific meta-regression within the CN subgroup did not find a significant association between mean cohort age and effect size (*p* = 0.221). This finding presents a conundrum at the aggregate level: while preclinical pathology is a plausible driver of heterogeneity, this macro-level analysis could not substantiate its link to age. This highlights the limitations of cohort-level data and motivates our subsequent biomarker-stratified longitudinal analysis to resolve this question mechanistically. Following our data processing protocol, only one cohort focused on Mild Cognitive Impairment (MCI) remained, which precluded a pooled subgroup estimate for this intermediate stage.

Further stratification revealed a clear gene-dose effect. Homozygous (*ε*4/*ε*4) carriers exhibited a large and significant reduction in hippocampal volume (SMD = -0.60, 95% CI [-1.08, -0.12], *p* = 0.015), whereas the effect in heterozygous (*ε*3/*ε*4) carriers was negligible and non-significant (SMD = -0.06, 95% CI [-0.31, 0.19], *p* = 0.652). Methodological factors also appeared to influence the results; studies that employed intracranial volume (ICV) correction reported a strong and significant effect (SMD = -0.41, 95% CI [-0.67, -0.16]), whereas uncorrected studies showed a null effect (SMD = - 0.10, 95% CI [-0.25, 0.06]).

To investigate sources of heterogeneity across the entire dataset, we performed univariable meta-regressions. We found that neither the mean age of participants (*p* = 0.71) nor the proportion of female participants (*p* = 0.57) were significant moderators of the overall effect size. Sensitivity analyses confirmed the stability of our findings; a multiverse robustness check, which involved sequentially removing the most extreme studies, showed that the overall conclusion remained unchanged. However, a trim-and-fill analysis suggested that smaller, non-significant studies might be underrepresented. The bias-corrected effect size was attenuated and no longer statistically significant (SMD = -0.10, 95% CI [-0.30, 0.10]), warranting caution in interpreting the precise magnitude of the overall effect and reinforcing the need for mechanistic validation in longitudinal cohorts.

### 3.2 NACC Cohort: Validation of Longitudinal Atrophy Rates

In the NACC longitudinal cohort, we analyzed 3,996 observations from 3,239 subjects using a linear mixed-effects model. The model was adjusted for baseline age, sex, eTIV, baseline clinical diagnosis, and their respective interactions with time where appropriate.

The analysis revealed a statistically significant baseline atrophy rate for non-carriers (the reference group), estimated at -31.61 mm^3^/year (*p* = 0.031). A significant interaction was found between time and the homozygous genotype, indicating an accelerated rate of atrophy in this group. Specifically, APOE-*ε*4 homozygotes showed an additional volume loss of 75.11 mm^3^/year relative to non-carriers (Interaction *β* = −75.11, *p* = 0.040). This corresponds to a total estimated annual atrophy rate of -106.72 mm^3^/year, which is approximately 3.4 times that of non-carriers (Figure 4).

**Figure 4.**
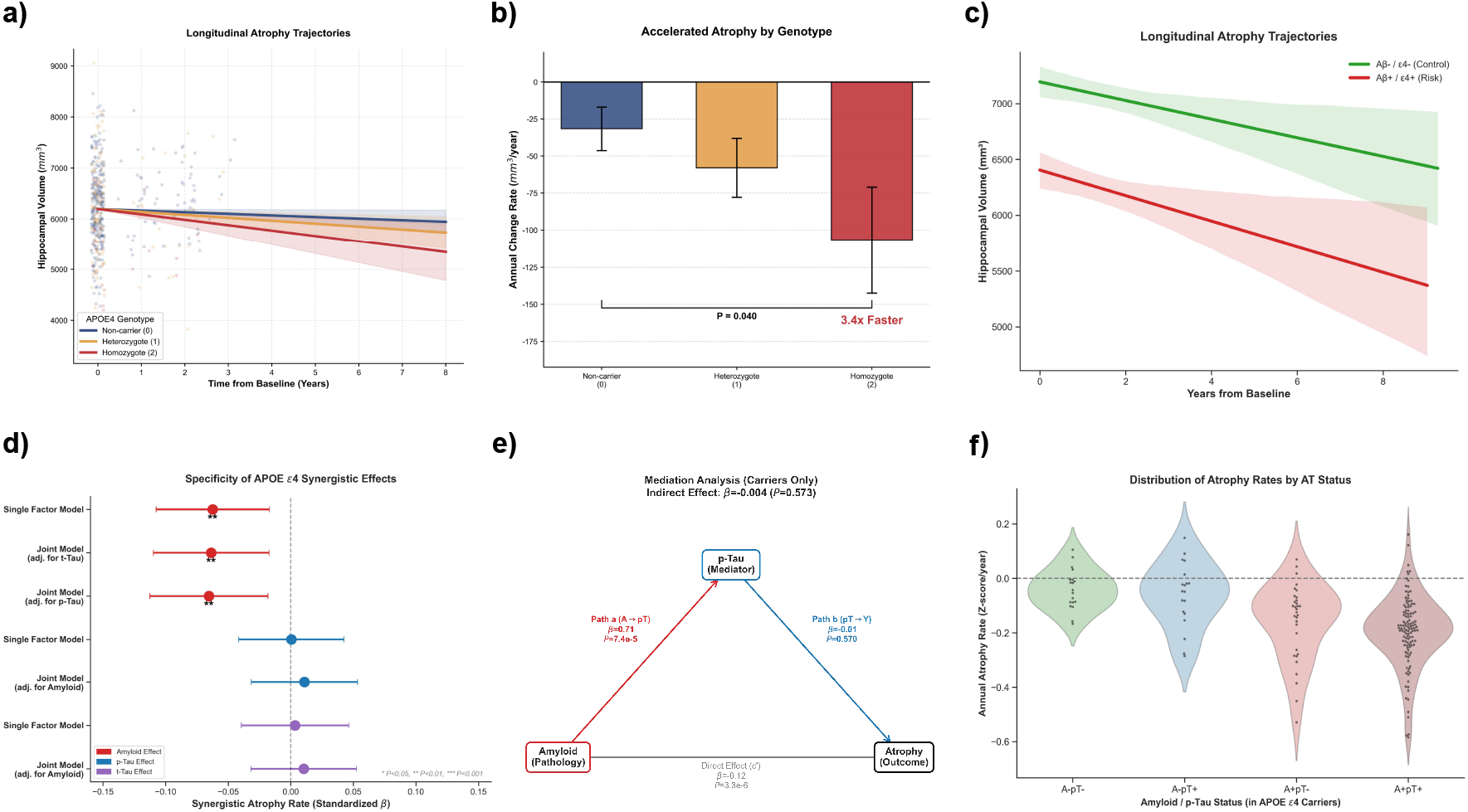
Longitudinal hippocampal atrophy trajectories and synergistic interaction effects in the NACC and ADNI cohorts. (a) Estimated longitudinal trajectories of hippocampal volume (mm^3^) in the NACC cohort, stratified by APOE-*ε*4 gene dosage (Non-carrier, Heterozygote, Homozygote). Shaded areas represent 95% Confidence Intervals (CIs). (b) Annualized atrophy rates (mm^3^/year) derived from the NACC Linear Mixed-Effects Model. Bars represent the additional atrophy rate associated with each genotype relative to the non-carrier reference group, quantified by the Time × Genotype interaction term. The model indicates a significantly accelerated decline in homozygotes (p=0.040). Error bars denote Standard Errors (SE). (c) Estimated longitudinal trajectories in the ADNI cohort, stratified by both Amyloid-*β* (A*β*) status and APOE-*ε*4 carrier status (A*β*+/*ε*4+ vs. the A*β*-/*ε*4- control group), illustrating the potent gene-pathology interaction. (d) Forest plot quantifying the specificity of the synergistic interaction effect (Time × APOE-*ε*4 × Biomarker) on atrophy rates from the ADNI LMMs. The amyloid interaction (red) is significant and remains robust in joint models adjusted for p-Tau or t-Tau, while p-Tau (blue) and t-Tau (purple) interactions are non-significant. Points represent standardized *β*-coefficients; horizontal lines are 95% CIs. (e) Path diagram of the mediation analysis in APOE-*ε*4 carriers, testing if the effect of A*β* pathology on atrophy is mediated by p-Tau. The analysis reveals a non-significant indirect effect (p=0.573), supporting a direct association between A*β* and atrophy. (f) Violin plots showing the distribution of annualized atrophy rates (Z-score/year) among APOE-*ε*4 carriers stratified by their AT status. Individuals positive for both amyloid and p-Tau (A+pT+) exhibit the fastest rates of neurodegeneration.

In contrast, for APOE-*ε*4 heterozygotes, the interaction term indicated a non-significant additional decline of - 26.37 mm^3^/year (*p* = 0.223). These results from a large, independent clinical cohort support the findings from our meta-analysis. They suggest a non-linear gene-dose effect, wherein the acceleration in hippocampal decline appears to be predominantly attributable to the homozygous state.

### 3.3 ADNI Cohort: Pathological Associations and Mechanisms of APOE-*ε*4 Synergism

Following quality control of the ADNI cohort data (final sample: 1,947 observations from 1,150 subjects), we first investigated whether the effect of APOE-*ε*4 on accelerating hippocampal atrophy was associated with specific pathological pathways. Results from individual LMM showed a statistically significant synergistic interaction between APOE-*ε*4 and A*β* positivity; their combination was associated with a faster rate of hippocampal atrophy (standardized *β* = −0.07, *P <* 0.01). In contrast, synergistic interactions between APOE-*ε*4 and either p-Tau (*P* = 0.57) or t-Tau (*P* = 0.62) did not reach statistical significance. To further assess the potential primary role of the A*β* pathway, we constructed a joint model that included interaction terms for both A*β* and p-Tau. In this model, the synergistic interaction between A*β* and APOE-*ε*4 remained significant even after adjusting for the influence of p-Tau (*β* = −0.07, *P <* 0.01), whereas the p-Tau interaction remained non-significant (*P >* 0.5). These results collectively suggest that the accelerating effect of APOE-*ε*4 on neurodegeneration appears to be primarily associated with the amyloid pathway (Table 2) (Figure 4).

**Table 2.**
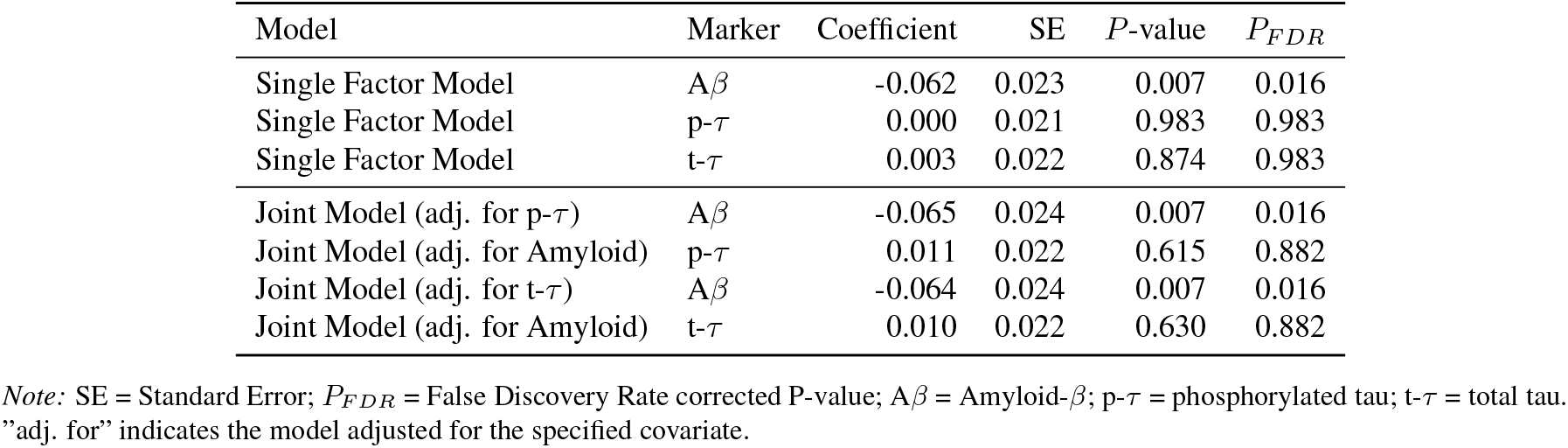
Statistical Model Results for Biomarkers.

This synergistic relationship between genotype and pathology was accompanied by observable differences in clinical trajectories. Compared to A*β*-negative non-carriers (A*β*-/*ε*4-), the A*β*-positive APOE-*ε*4 carrier group (A*β*+/*ε*4+) exhibited a steeper decline in hippocampal volume over the longitudinal follow-up.

Based on these findings, we next performed a mediation analysis to investigate the intrinsic relationship among A*β*, p-Tau, and brain atrophy within APOE-*ε*4 carriers. The analysis indicated that A*β* pathology was a strong predictor of elevated p-Tau levels (Path a: *β* = 0.710, *P <* 0.001), but p-Tau level was not significantly associated with the rate of atrophy after accounting for the direct effect of A*β* (Path b: *β* = −0.010, *P* = 0.570). Consequently, the test results did not support an indirect pathway from A*β* to atrophy via p-Tau (Indirect Effect : *β* = −0.004, *P* = 0.573). At the same time, the direct effect of A*β* on atrophy remained significant (Direct Effect : *β* = −0.120, *P <* 0.001). These findings provide evidence for the view that, in APOE-*ε*4 carriers, the A*β*-related acceleration of atrophy is consistent with a direct pathway rather than being primarily mediated by p-Tau.

Although p-Tau does not appear to be a mediator in this pathway, its presence remains an indicator of disease severity. After stratifying APOE-*ε*4 carriers by their AT status, it was observed that individuals positive for both A*β* and p-Tau (A+pT+) exhibited the fastest mean rate of atrophy. This suggests that when p-Tau pathology co-occurs with A*β* pathology and genetic risk, neurodegeneration may be in its most aggressive phase.

## 4 Discussion

This study integrates multi-cohort evidence to address conflicting reports regarding the impact of APOE-*ε*4 on hippocampal integrity. Our findings support a perspective where APOE-*ε*4 functions not as a static risk factor exerting a constant deleterious effect, but as a conditional modulator dependent on the presence of amyloid-*β*.

The heterogeneity identified in our meta-analysis highlights the complexity of detecting this effect in aggregate data. Notably, while we hypothesized that older age, serving as a proxy for increased occult amyloid prevalence, would correlate with larger effect sizes in Cognitively Normal cohorts, our meta-regression did not support this association (*p* = 0.221). This discrepancy between macro-level aggregate data and the expected biological trend likely stems from the ecological fallacy, where mean cohort characteristics obscure individual-level heterogeneity. Consequently, the inability of simple demographic factors to predict atrophy risk in the meta-analysis underscores the critical necessity of our subsequent longitudinal validation. By transitioning from cohort-level aggregates to individual-level biomarker stratification, we resolved this apparent contradiction. We demonstrated that in the absence of A*β*, even older APOE-*ε*4 carriers exhibit atrophy trajectories statistically indistinguishable from non-carriers. This confirms that variable findings in prior literature likely reflect differing prevalences of occult amyloidosis that cannot be captured by age demographics alone.This observation serves as a cautionary note for future meta-analytic studies, warning against reliance on aggregate demographic data and emphasizing the critical need for biomarker-level stratification to achieve mechanistic clarity.

Our analysis of the NACC cohort reveals a crucial non-linearity in the allele dosage effect. We observed a step-wise increase in hippocampal loss characterized by a distinct threshold: homozygous (*ε*4/*ε*4) carriers exhibited a marked acceleration in atrophy, approximately three-fold that of non-carriers, whereas the effect in heterozygotes was minimal and not statistically significant. This suggests that the association of APOE with neurodegeneration is cumulative and threshold-dependent, indicating that homozygosity represents a distinct state of elevated biological risk [54, 55].

Mechanistically, our ADNI results provide insight into the drivers of this acceleration. We identified a robust synergistic interaction between APOE-*ε*4 and A*β* positivity. The mediation analysis indicated that while A*β* strongly predicts p-Tau levels, the accelerated atrophy in carriers was associated directly with A*β*, with no statistically significant indirect effect via soluble p-Tau (p-Tau181). However, this finding warrants cautious interpretation, particularly as our analysis was based on a soluble fluid biomarker. It does not necessarily imply that Tau pathology is biologically irrelevant or absent [56]. Rather, it suggests that in the specific context of APOE-*ε*4, the neurodegenerative cascade may not rely exclusively on the sequential amyloid-tau-neurodegeneration pathway captured by fluid biomarkers. Future studies using Tau-PET imaging will be essential to definitively clarify this mediation pathway, as fluid biomarkers may lag behind or behave differently from the aggregated neurofibrillary tangles more proximal to neurodegeneration. A plausible explanation for this direct link involves the innate immune response. Emerging evidence suggests that APOE-*ε*4 may impair the ability of microglia to contain amyloid plaques, shifting them towards a neurodegenerative phenotype that promotes tissue loss through neuroinflammatory mechanisms [9, 57]. These pathways might operate in parallel with, or precede, extensive neurofibrillary tangle formation. Thus, APOE-*ε*4 may amplify the susceptibility of the brain to amyloid toxicity through pleiotropic mechanisms, creating a scenario where amyloid presence serves as a sufficient trigger for accelerated structural loss [13].

The amyloid-dependent and dose-sensitive relationship described here has implications for disease-modifying therapies. If the structural risk associated with APOE-*ε*4 is conditional on amyloid burden, therapeutic plaque clearance could theoretically attenuate the acceleration of atrophy [58, 59]. Our results suggest this clinical benefit might be most critical for homozygous carriers, who face the steepest natural trajectory of decline, although potential confounders such as amyloid-related imaging abnormalities (ARIA) must be considered [60].

Several limitations should be considered. First, the inability of the meta-regression to detect age effects highlights the noise inherent in aggregating diverse methodological protocols. Second, while we utilized p-Tau181 as a marker, it primarily reflects soluble phosphorylated tau and may not fully capture the burden of insoluble neurofibrillary tangles, potentially underestimating the role of Tau in our mediation model. Future confirmation with Tau-PET imaging will therefore be crucial to investigate the role of aggregated tau pathology in this pathway. Third, the longitudinal cohorts are subject to selection bias, likely representing a healthier subset of the elderly population compared to the general community. Finally, consistent post-processing protocols were prioritized to mitigate measurement variations across scanner versions [61].

## 5 Conclusion

This study characterizes APOE-*ε*4 as a conditional, dose-dependent accelerator of hippocampal atrophy associated with amyloid pathology. We demonstrate that the limitation of macro-level demographics to predict atrophy risk is resolved by granular, individual-level biomarker profiling. The observed acceleration in homozygous carriers appears driven by a synergistic interaction with amyloid. While our statistical models point to a direct association between A*β* and atrophy in carriers, this likely reflects complex, pleiotropic mechanisms, potentially involving neuroinflammation, where APOE-*ε*4 exacerbates the neurotoxic response to amyloid. Consequently, clinical risk stratification should integrate amyloid status with allele dosage, and homozygous carriers may represent a priority population for early antiamyloid interventions to mitigate irreversible structural loss.

## Acknowledgments

The authors extend their sincere gratitude to their colleagues and mentors, as well as to Shenzhen X-Institute and Xinjiang University, for their invaluable discussions and support.

Data collection and sharing for the Alzheimer’s Disease Neuroimaging Initiative (ADNI) is funded by the National Institute on Aging (National Institutes of Health Grant U19AG024904). The grantee organization is the Northern California Institute for Research and Education. In the past, ADNI has also received funding from the National Institute of Biomedical Imaging and Bioengineering, the Canadian Institutes of Health Research, and private sector contributions through the Foundation for the National Institutes of Health (FNIH) including generous contributions from the following: AbbVie, Alzheimer’s Association; Alzheimer’s Drug Discovery Foundation; Araclon Biotech; BioClinica, Inc.; Biogen; Bristol-Myers Squibb Company; CereSpir, Inc.; Cogstate; Eisai Inc.; Elan Pharmaceuticals, Inc.; Eli Lilly and Company; EuroImmun; F. Hoffmann-La Roche Ltd and its affiliated company Genentech, Inc.; Fujirebio; GE Healthcare; IXICO Ltd.; Janssen Alzheimer Immunotherapy Research & Development, LLC.; Johnson & Johnson Pharmaceutical Research & Development LLC.; Lumosity; Lundbeck; Merck & Co., Inc.; Meso Scale Diagnostics, LLC.; NeuroRx Research; Neurotrack Technologies; Novartis Pharmaceuticals Corporation; Pfizer Inc.; Piramal Imaging; Servier; Takeda Pharmaceutical Company; and Transition Therapeutics.

The NACC database is funded by NIA/NIH Grant U24 AG072122. SCAN is a multi-institutional project that was funded as a U24 grant (AG067418) by the National Institute on Aging in May 2020. Data collected by SCAN and shared by NACC are contributed by the NIA-funded ADRCs as follows: Arizona Alzheimer’s Center - P30 AG072980 (PI: Eric Reiman, MD); R01 AG069453 (PI: Eric Reiman (contact), MD); P30 AG019610 (PI: Eric Reiman, MD); and the State of Arizona which provided additional funding supporting our center; Boston University - P30 AG013846 (PI Neil Kowall MD); Cleveland ADRC - P30 AG062428 (James Leverenz, MD); Cleveland Clinic, Las Vegas – P20AG068053; Columbia - P50 AG008702 (PI Scott Small MD); Duke/UNC ADRC – P30 AG072958; Emory University - P30AG066511 (PI Levey Allan, MD, PhD); Indiana University - R01 AG19771 (PI Andrew Saykin, PsyD); P30 AG10133 (PI Andrew Saykin, PsyD); P30 AG072976 (PI Andrew Saykin, PsyD); R01 AG061788 (PI Shannon Risacher, PhD); R01 AG053993 (PI Yu-Chien Wu, MD, PhD); U01 AG057195 (PI Liana Apostolova, MD); U19 AG063911 (PI Bradley Boeve, MD); and the Indiana University Department of Radiology and Imaging Sciences; Johns Hopkins - P30 AG066507 (PI Marilyn Albert, Phd.); Mayo Clinic - P50 AG016574 (PI Ronald Petersen MD PhD); Mount Sinai - P30 AG066514 (PI Mary Sano, PhD); R01 AG054110 (PI Trey Hedden, PhD); R01 AG053509 (PI Trey Hedden, PhD); New York University - P30AG066512-01S2 (PI Thomas Wisniewski, MD); R01AG056031 (PI Ricardo Osorio, MD); R01AG056531 (PIs Ricardo Osorio, MD; Girardin Jean-Louis, PhD); Northwestern University - P30 AG013854 (PI Robert Vassar PhD); R01 AG045571 (PI Emily Rogalski, PhD); R56 AG045571, (PI Emily Rogalski, PhD); R01 AG067781, (PI Emily Rogalski, PhD); U19 AG073153, (PI Emily Rogalski, PhD); R01 DC008552, (M.-Marsel Mesulam, MD); R01 AG077444, (PIs M.-Marsel Mesulam, MD, Emily Rogalski, PhD); R01 NS075075 (PI Emily Rogalski, PhD); R01 AG056258 (PI Emily Rogalski, PhD); Oregon Health & Science University - P30 AG066518 (PI Lisa Silbert, MD, MCR); Rush University - P30 AG010161 (PI David Bennett MD); Stanford – P30AG066515; P50 AG047366 (PI Victor Henderson MD MS); University of Alabama, Birmingham – P20; University of California, Davis - P30 AG10129 (PI Charles DeCarli, MD); P30 AG072972 (PI Charles DeCarli, MD); University of California, Irvine - P50 AG016573 (PI Frank LaFerla PhD); University of California, San Diego - P30AG062429 (PI James Brewer, MD, PhD); University of California, San Francisco - P30 AG062422 (Rabinovici, Gil D., MD); University of Kansas - P30 AG035982 (Russell Swerdlow, MD); University of Kentucky - P30 AG028283-15S1 (PIs Linda Van Eldik, PhD and Brian Gold, PhD); University of Michigan ADRC - P30AG053760 (PI Henry Paulson, MD, PhD) P30AG072931 (PI Henry Paulson, MD, PhD) Cure Alzheimer’s Fund 200775 - (PI Henry Paulson, MD, PhD) U19 NS120384 (PI Charles DeCarli, MD, University of Michigan Site PI Henry Paulson, MD, PhD) R01 AG068338 (MPI Bruno Giordani, PhD, Carol Persad, PhD, Yi Murphey, PhD) S10OD026738-01 (PI Douglas Noll, PhD) R01 AG058724 (PI Benjamin Hampstead, PhD) R35 AG072262 (PI Benjamin Hampstead, PhD) W81XWH2110743 (PI Benjamin Hampstead, PhD) R01 AG073235 (PI Nancy Chiaravalloti, University of Michigan Site PI Benjamin Hampstead, PhD) 1I01RX001534 (PI Benjamin Hampstead, PhD) IRX001381 (PI Benjamin Hampstead, PhD); University of New Mexico - P20 AG068077 (Gary Rosenberg, MD); University of Pennsylvania - State of PA project 2019NF4100087335 (PI David Wolk, MD); Rooney Family Research Fund (PI David Wolk, MD); R01 AG055005 (PI David Wolk, MD); University of Pittsburgh - P50 AG005133 (PI Oscar Lopez MD); University of Southern California - P50 AG005142 (PI Helena Chui MD); University of Washington - P50 AG005136 (PI Thomas Grabowski MD); University of Wisconsin - P50 AG033514 (PI Sanjay Asthana MD FRCP); Vanderbilt University – P20 AG068082; Wake Forest - P30AG072947 (PI Suzanne Craft, PhD); Washington University, St. Louis - P01 AG03991 (PI John Morris MD); P01 AG026276 (PI John Morris MD); P20 MH071616 (PI Dan Marcus); P30 AG066444 (PI John Morris MD); P30 NS098577 (PI Dan Marcus); R01 AG021910 (PI Randy Buckner); R01 AG043434 (PI Catherine Roe); R01 EB009352 (PI Dan Marcus); UL1 TR000448 (PI Brad Evanoff); U24 RR021382 (PI Bruce Rosen); Avid Radiopharmaceuticals / Eli Lilly; Yale - P50 AG047270 (PI Stephen Strittmatter MD PhD); R01AG052560 (MPI: Christopher van Dyck, MD; Richard Carson, PhD); R01AG062276 (PI: Christopher van Dyck, MD); 1Florida - P30AG066506-03 (PI Glenn Smith, PhD); P50 AG047266 (PI Todd Golde MD PhD).

The Alzheimer’s Disease Genetics Consortium (ADGC) is funded by a grant from the National Institute on Aging (PI, Gerard D. Schellenberg; UO1AG032984).

## Code Availability

The analysis code for this paper is available at the GitHub repository: https://github.com/wyqmath/admeta.

## A Supporting Information

### A.1 Search Strategy and Study Selection

A comprehensive literature search was conducted in four electronic databases from their inception to August 2025: PubMed, Embase (via Ovid), the Cochrane Library, and Web of Science (Science Citation Index Expanded and Emerging Sources Citation Index). The search was limited to publications in English. The search strategy combined subject headings (MeSH in PubMed, Emtree in Embase) with free-text keywords covering two core concepts: (1) Alzheimer’s disease and related terms; and (2) Apolipoprotein E *ϵ*4 (APOE *ϵ*4) carrier status. No methodological filters for study design were applied during the search phase to maximize sensitivity. The complete search queries for all databases are detailed in Supplementary Table S1.

All retrieved records were first imported into EndNote for reference management and automated deduplication. Subsequently, the screening process was conducted in two stages by two independent researchers. In the first stage, titles and abstracts were screened to exclude reviews, metaanalyses, animal studies, letters, case reports, conference proceedings, and literature clearly irrelevant to the research topic. In the second stage, the remaining records were uploaded to the web-based systematic review platform Covidence (https://www.covidence.org/) for full-text assessment. These two researchers independently reviewed the full texts against specific eligibility criteria, which are detailed in the following section. To ensure the independence of data, we rigorously checked publications to identify potential population overlap. When multiple articles reported on the same or highly similar datasets, we included only the one with the largest sample size or the most recent publication date. Any discrepancies during the screening process were resolved through discussion or by a third researcher’s adjudication. The entire screening process is reported in a PRISMA 2020 flow diagram.

### A.2 Data Extraction

Two researchers independently extracted data from the included studies using a pre-designed standardized data extraction form. Any disagreements were resolved by consulting a third researcher. We extracted the following demographic and clinical characteristics: sample size, mean age, sex ratio, and APOE *ϵ*4 gene dosage information for each experimental group. The primary outcome measures extracted included total hippocampal volume, as well as left and right hippocampal volumes separately. For studies that only reported left and right hippocampal volumes without the total volume, we used a standard formula incorporating the correlation coefficient to calculate the mean and standard deviation of the total hippocampal volume, correcting for the correlation between the two hemispheres. To address potential methodological heterogeneity, we also recorded detailed information regarding MRI image acquisition and processing. This included: (1) the specific MRI processing software used; (2) the hippocampal segmentation method (manual tracing vs. automated segmentation); (3) whether hippocampal volumes were corrected for intracranial volume (ICV) to account for individual differences in head size, and the specific correction method used; and (4) whether longitudinal data processing was included.

### A.3 Eligibility Criteria

The inclusion and exclusion criteria were established *a priori* based on the PICOS (Population, Intervention/Exposure, Comparator, Outcomes, Study design) framework and reported according to the PRISMA guidelines.

#### A.3.1 Inclusion Criteria

Studies were considered eligible for inclusion if they met the following criteria:

##### 1. Study Design

The study design was cross-sectional or provided baseline data from longitudinal cohorts comparing *APOE ϵ*4 carriers versus non-carriers.

##### 2. Population

Participants were human adults diagnosed with Alzheimer’s disease (AD), Mild Cognitive Impairment (MCI), or were Cognitively Normal (CN) based on established clinical criteria (e.g., NINCDS-ADRDA, NIA-AA).

##### 3. Outcome

The study reported quantitative measurements of hippocampal volume derived from Magnetic Resonance Imaging (MRI), regardless of the segmentation method (manual or automated) or intracranial volume (ICV) correction status.

##### 4. Data Availability

Sufficient statistical data were provided, specifically the sample size (*N*), mean hippocampal volume, and standard deviation (*SD*) or standard error (*SE*) for both genotype groups to allow for the calculation of effect sizes.

##### 5. Publication Type

The article was an original research paper published in English.

#### A3.2. Exclusion Criteria

Studies were excluded based on the following criteria:

##### 1. Non-Original Research

Non-original research, including reviews, meta-analyses, conference abstracts, letters, editorials, or case reports.

##### 2. Participants

Studies involving animal models or *in vitro* experiments, or studies focusing on participants with other neurological or psychiatric disorders (e.g., Parkinson’s disease, vascular dementia) that could confound hippocampal atrophy assessments.

##### 3. Data Quality

Publications with insufficient or irretrievable data where the Standardized Mean Difference (*SMD*) could not be calculated.

##### 4. Data Overlap

Studies utilizing overlapping datasets or cohorts. In cases where multiple studies reported on the same population (e.g., overlapping samples from the ADNI database), priority was given to the study with the largest sample size or the most recent publication date to avoid data duplication.

---

**Table S1.** Detailed Search Strategies for Electronic Databases (Search Date: August 2025)

| Database | Search Strategy |
| --- | --- |
| **1. PubMed** |  |
| #1 | "Alzheimer Disease"[Mesh] OR "Alzheimer*"[Title/Abstract] OR "Alzheimer's"[Title/Abstract] OR "AD"[Title/Abstract] OR "Senile Dementia"[Title/Abstract] OR "Presenile Dementia"[Title/Abstract] OR "Alzheimer Type Dementia"[Title/Abstract] OR "FAD"[Title/Abstract] OR "Familial Alzheimer Disease"[Title/Abstract] OR "Early Onset Alzheimer*"[Title/Abstract] OR "Late Onset Alzheimer*"[Title/Abstract] |
| #2 | "Apolipoprotein E4"[Mesh] OR "Apolipoprotein E"[Mesh] OR "APOE"[Title/Abstract] OR "APOE4"[Title/Abstract] OR "ApoE-4"[Title/Abstract] OR "epsilon 4"[Title/Abstract] OR "e4 allele"[Title/Abstract] OR "Apo E"[Title/Abstract] |
| #3 | "Hippocampus"[Mesh] OR "Hippocamp*"[Title/Abstract] OR "Hippocampal Volume"[Title/Abstract] OR "Hippocampal Atrophy"[Title/Abstract] |
| #4 | "Magnetic Resonance Imaging"[Mesh] OR "MRI"[Title/Abstract] OR "Neuroimaging"[Title/Abstract] OR "Voxel-based morphometry"[Title/Abstract] OR "Volumetric"[Title/Abstract] OR "Brain Volume"[Title/Abstract] |
| #5 | #3 AND #4 |
| #6 | #1 AND #2 AND #5 |
| **2. Embase** |  |
| #1 | 'alzheimer disease'/exp OR 'alzheimer*':ti,ab,kw OR 'senile dementia':ti,ab,kw OR 'presenile dementia':ti,ab,kw OR 'ad':ti,ab,kw OR 'familial alzheimer disease':ti,ab,kw |
| #2 | 'apolipoprotein E4'/exp OR 'apoe':ti,ab,kw OR 'apoe4':ti,ab,kw OR 'apolipoprotein e':ti,ab,kw OR 'epsilon 4':ti,ab,kw |
| #3 | 'hippocampus'/exp OR 'hippocamp*':ti,ab,kw OR 'hippocampal atrophy':ti,ab,kw OR 'hippocampal volume':ti,ab,kw |
| #4 | 'nuclear magnetic resonance imaging'/exp OR 'mri':ti,ab,kw OR 'magnetic resonance imaging':ti,ab,kw |
| #5 | #1 AND #2 AND #3 AND #4 |
| **3. Web of Science** |  |
| #1 | TS=("Alzheimer*" OR "AD" OR "Senile Dementia" OR "Presenile Dementia" OR "Alzheimer Type Dementia" OR "FAD") |
| #2 | TS=("Apolipoprotein E" OR "APOE" OR "APOE4" OR "epsilon 4" OR "Apo E4") |
| #3 | TS=("Hippocampus" OR "Hippocampal" OR "Hippocampal Volume" OR "Hippocampal Atrophy") |
| #4 | TS=("MRI" OR "Magnetic Resonance Imaging" OR "Brain Volume") |
| #5 | #1 AND #2 AND #3 AND #4 |
| **4. Cochrane Library** |  |
| #1 | MeSH descriptor: [Alzheimer Disease] explode all trees |
| #2 | (Alzheimer* OR "Senile Dementia" OR "Presenile Dementia" OR "Alzheimer Type Dementia" OR "FAD" OR "Familial Alzheimer Disease"):ti,ab,kw |
| #3 | #1 OR #2 |
| #4 | MeSH descriptor: [Apolipoprotein E4] explode all trees |
| #5 | MeSH descriptor: [Apolipoproteins E] explode all trees |
| #6 | (APOE* OR "Apolipoprotein E" OR "Apolipoprotein E4" OR "epsilon 4" OR "e4 allele"):ti,ab,kw |
| #7 | #4 OR #5 OR #6 |
| #8 | MeSH descriptor: [Hippocampus] explode all trees |
| #9 | (Hippocamp* OR "Hippocampal Volume" OR "Hippocampal Atrophy"):ti,ab,kw |
| #10 | #8 OR #9 |
| #11 | MeSH descriptor: [Magnetic Resonance Imaging] explode all trees |
| #12 | (MRI OR "Magnetic Resonance Imaging" OR "Brain Volume" OR Volumetric OR "Voxel-based morphometry"):ti,ab,kw |
| #13 | #11 OR #12 |
| #14 | #3 AND #7 AND #10 AND #13 |

**Table S2.** Characteristics of Included Studies for Meta-Analysis.

| Study ID | Sample Size | Age (Mean ± SD) | Gender (% Male) | Included Diagnosis |
| --- | --- | --- | --- | --- |
| [20] Khan et al., 2017 | 1781 | 74.4 (Weighted Mean) | 49.6 | AD, MCI, CN |
| [21] Pievani et al., 2011 | 56 | 71.1 ± 8.3 | 32.1 | AD, CN |
| [22] Geroldi et al., 1999 | 58 | 70.7 ± 8.7 | 27.6 | AD, CN |
| [23] Plassman et al., 1997 | 20 | 62.5 ± 7.8 | 30 | CN |
| [24] Adamson et al., 2010 | 50 | 61.0 ± 6.7 | 82 | CN |
| [42] Dong et al., 2019 | 117 | 58.1 ± 6.1 | 38.5 | CN |
| [25] Koivumäki et al., 2024 | 59 | 67.6 ± 4.7 | 36.7 | CN |
| [26] Soininen et al., 1995 | 32 | 69 ± 6 | 31.3 | CN |
| [27] Hostage et al., 2013 | 662 | 75.3 ± 6.6 | 58.2 | AD, MCI, CN |
| [28] Bussy et al., 2019 | 162 | 38.8 ± 11.7 | 39.5 | CN |
| [29] Chang et al., 2019 | 97 | 71.5 ± 7.8 | 54.6 | AD |
| [30] Alexopoulos et al., 2011 | 33 | 24.5 ± 3.6 | 36.4 | CN |
| [31] Reiman et al., 1998 | 33 | 50–62 (Range) | NR | CN |
| [32] Den Heijer et al., 2002 | 428 | 72.3 ± 7.0 | 47.8 | CN |
| [33] Chiang et al., 2011 | 297 | 74.9 ± 7.2 | 59.9 | AD, MCI, CN |
| [34] Westlye et al., 2011 | 95 | 63.8 ± 7.2 | 35.8 | CN |
| [35] Richter-Schmidinger et al., 2011 | 135 | 24.5 ± 3.2 | 32.6 | CN |
| [36] An et al., 2016 | 637 | 72.3 ± 7.4 | 58.4 | MCI |
| [6] O'Dwyer et al., 2012 | 44 | 26.8 ± 4.6 | 59.1 | CN |
| [37] Lind et al., 2006 | 60 | 66.0 ± 8.1 | 38.3 | CN |
| [38] Wang et al., 2019 | 1347 | 73.6 ± 7.1 | 55.8 | AD, MCI, CN |
| [39] Lehtovirta et al., 1995 | 42 | 69.4 ± 7.5 | 47.6 | AD, CN |
| [40] Lemaître et al., 2005 | 750 | 69.4 ± 2.9 | 42 | CN |
| [41] Cherbuin et al., 2008 | 331 | 62.6 ± 1.4 | 53.5 | CN |

**Table S3.** Full Results of the Linear Mixed-Effects Model on NACC Data.

| Predictor | Coefficient | Std. Error | z-value | P-value | 95% Confidence Interval |
| --- | --- | --- | --- | --- | --- |
| *Fixed Effects* |  |  |  |  |  |
| Intercept | 3693.02 | 101.95 | 36.22 | < 0.001 | [3493.20, 3892.84] |
| *APOE4 Dosage (Ref: Non-carrier)* |  |  |  |  |  |
| Heterozygote (1) | -98.29 | 25.02 | -3.93 | < 0.001 | [-147.33, -49.25] |
| Homozygote (2) | -342.35 | 51.31 | -6.67 | < 0.001 | [-442.91, -241.79] |
| Sex (Ref: Female) [Male] | 227.65 | 28.14 | 8.09 | < 0.001 | [172.50, 282.79] |
| *Baseline Diagnosis (Ref: CN)* |  |  |  |  |  |
| AD | -721.25 | 44.22 | -16.31 | < 0.001 | [-807.92, -634.57] |
| MCI | -284.91 | 28.33 | -10.06 | < 0.001 | [-340.43, -229.39] |
| Time | -31.61 | 14.68 | -2.15 | 0.031 | [-60.38, -2.84] |
| Time × APOE4 Heterozygote | -26.37 | 21.62 | -1.22 | 0.223 | [-68.76, 16.01] |
| Time × APOE4 Homozygote | -75.11 | 36.49 | -2.06 | 0.040 | [-146.63, -3.60] |
| Time × Sex [Male] | -28.40 | 19.91 | -1.43 | 0.154 | [-67.43, 10.63] |
| Age (Centered) | -23.98 | 1.38 | -17.33 | < 0.001 | [-26.69, -21.27] |
| Time × Age (Centered) | -3.44 | 1.25 | -2.75 | 0.006 | [-5.89, -0.99] |
| eTIV (Scaled) | 1.75 | 0.07 | 24.10 | < 0.001 | [1.61, 1.89] |
| *Random Effects Variance Components* |  |  |  |  |  |
| Group Variance | 15.79 |  |  |  |  |
| Group × Time Covariance | -0.81 |  |  |  |  |
| Time Variance | 1.65 |  |  |  |  |

**Figure S1.**
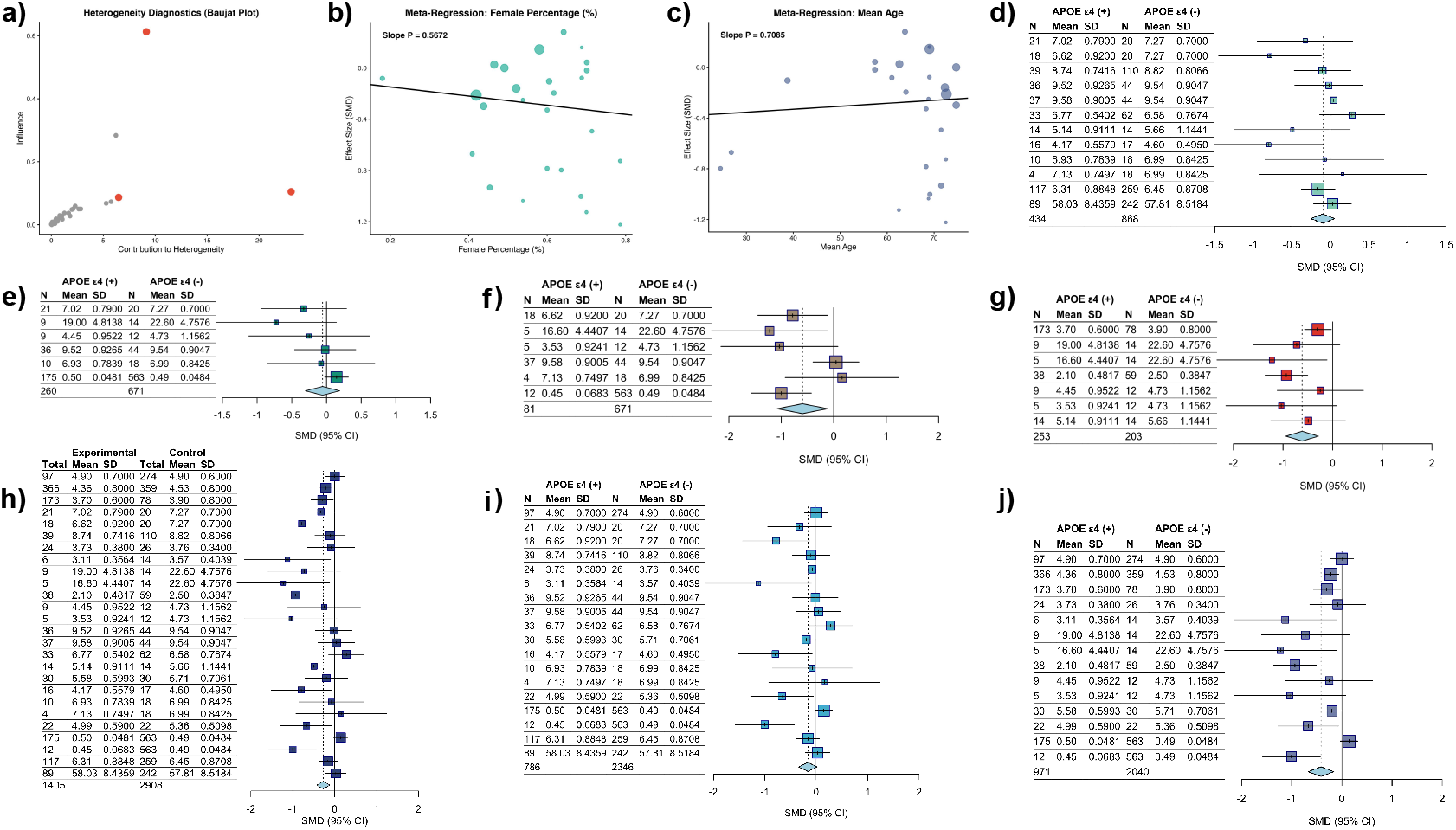
Supplementary plots for the meta-analysis. (a) Baujat plot for heterogeneity diagnostics, identifying influential studies. (b-c)Meta-regression plots showing the relationship between effect size (SMD) and moderators: (b) percentage of female participants and (c) mean age. (d-j) Forest plots visualizing the effect size of APOE *ϵ*4 on hippocampal volume across different subgroups and methodologies.

